# A Novel Collaborative Learning Model for Teeth and Fillings in Radiographs

**DOI:** 10.1101/2023.03.01.23286626

**Authors:** Erin Ealba Bumann, Saeed Al-Qarni, Geetha Chandrashekar, Roya Sabzian, Brenda Bohaty, Yugyung Lee

## Abstract

It is critical for dentists to identify and differentiate primary and permanent teeth, fillings, dental restorations and areas with pathological findings when reviewing dental radiographs to ensure that an accurate diagnosis is made and the optimal treatment can be planned. Unfortunately, dental radiographs are sometimes read incorrectly due to human error or low-quality images. While secondary or group review can help catch errors, many dentists work in practice alone and/or do not have time to review all of their patients’ radiographs with another dentist. Artificial intelligence may facilitate the accurate interpretation of radiographs. To help support the review of panoramic radiographs, we developed a novel collaborative learning model that simultaneously identifies and differentiates primary and permanent teeth and detects fillings. We used publicly accessible dental panoramic radiographic images and images obtained from the University of Missouri-Kansas City School of Dentistry to develop and optimize two high-performance classifiers: (1) a system for tooth segmentation that can differentiate primary and permanent teeth and (2) a system to detect dental fillings. By utilizing these high-performance classifiers, we created models that can identify primary and permanent teeth, as well as their associated dental fillings. We also designed a novel method for collaborative learning that utilizes these two classifiers to enhance recognition performance. Our model improves upon the existing machine learning models to simultaneously identify and differentiate primary and permanent teeth, and to identify any associated fillings.

## Introduction

It is crucial for dentists to be able to precisely identify all primary teeth, permanent teeth, and fillings, in addition to other types of dental restorations and pathologies, when reviewing dental radiographs. An incorrect interpretation of radiographs can lead to wasted time for both the dentist and patient due to the need for repeated appointments, or can potentially result in an inappropriate treatment for a patient. Unfortunately, human error can complicate image review. While secondary review by other dentist(s) in the practice can help reduce errors and provide a consensus interpretation of less-than-perfect images, many dentists work independently, and even those in large practices may not have time to obtain a second review for all patients.

Artificial intelligence and machine learning (AI/ML) methods are already making a major impact in healthcare by changing how images are reviewed and analyzed, diagnoses are made, and procedures are planned (Lee et al. 2022, Malamateniou et al. 2021, Song et al. 2022). Moving this image processing capability into the field of dentistry can improve the ability to diagnose and treat oral, dental, and craniofacial conditions. Numerous studies suggest that an autonomous system that can accurately detect teeth and dental restorations can be useful to objectively assess an individual’s oral health and plan an optimal course of treatment (Bayrakdar et al. 2022, Carillo-Perez et al. 2022, Gurses and Oktay 2020, Revilla-León 2022, Schwendicke et al. 2019).

Many distinct tooth segmentation techniques (partitioning an image to identify specific teeth) have been developed. Almost every model reported had an accuracy of at least 75% (compared to an expert’s classification), with most having accuracies >95% (Estai et al. 2022, Leite et al. 2021, Shaheen et al. 2021, Vinayahalingam et al. 2021). For example, Zhao et al. created a two-staged attention segmentation network (TSASNet) to localize and classify teeth in radiographs, which had an accuracy of approximately 97% (Zhao et al. 2020). Another group introduced a different two-staged network architecture, ToothNet, which utilized a supervised deep learning approach to capture the edge map from cone beam computed tomography (CBCT) images, then a region proposal network (RPN) to autonomously segment teeth (Cui et al. 2019). Jader and colleagues investigated deep learning approaches and chose a mask region-based convolutional neural network (CNN) (Mask R-CNN) that was able to segment every tooth, even in difficult panoramic radiographs (Jader et al. 2018). Another deep learning solution for automatic tooth segmentation based on panoramic radiographs was developed using the Mask R-CNN algorithm and annotated datasets (Lee et al. 2020).

Wirtz et al. provided a coupled shaped model for robust and accurate tooth segmentation in low-quality panoramic radiographs (Wirtz et al. 2018). Their model employed a deep neural network to obtain the binary mask of the teeth to statistically identify form and space changes and thereby improve the segmentation quality. A new method was pioneered for segmenting teeth, including tooth compartments (pulp, enamel), by combining a marker-controlled watershed (MCW) algorithm with local threshold techniques to assess CBCT images (Kakehbaraei et al. 2018). Zhang et al. proposed a model that used a deep CNN for accurate and autonomous segmentation, with an average accuracy of 98.8%, which was considered to be suitable for dental computer-aided design (CAD) systems (Zhang et al. 2020). Another 3-D system (the TSegNet approach) for tooth segregation was evaluated by Cui et al., and showed faster and more accurate segmentation, even accounting for uncertainties caused by missing, crowded, or misaligned teeth (Cui et al. 2021).

An analysis by Silva et al. compared different neural networks, including Mask R-CNN, hybrid task cascade (HTC), PANet, and ResNet, to perform tooth numbering and segmentation on difficult dental radiographs (Silva et al. 2020). Their results indicated that, while all frameworks could be used to estimate the size, number, and localization of teeth, the accuracy varied greatly, and was reduced when teeth were damaged or otherwise altered. Thus, while some of the reported methods of tooth segmentation show good accuracy under certain conditions, additional work is still needed.

There have also been a few previous studies that examined the primary dentition. One focused solely on segmenting out the primary dentition (Kılıc et al. 2021). Another segmented out both the primary and permanent dentition, but had a low mean average precision (mAP)(Pinheiro et al. 2021). Several studies have evaluated the use of machine learning approaches to identify mesiodens (supernumery teeth) in primary, mixed and permanent dentition, with accuracies ranging from ∼87% to ∼98% (Ha et al. 2021, Jeon et al. 2022, Ahn et al. 2021). A recent study used CNN to detect and number both primary and permanent teeth, with relatively high accuracy (Kaya 2022). Another classified patients into those with 32 or more teeth and those with fewer than 32 teeth, and found that the segmentation accuracy was higher for patients with 32 or more teeth, likely due to the greater consistency in the locations of the teeth (Kanuri et al. 2022). Although their model was able to detect teeth even in blurred radiographs, better models are needed to detect both primary and mixed (primary + permanent) dentition in panoramic radiographs, and to simultaneously detect both normal teeth and teeth with pathological findings or restorations.

We herein describe a novel collaborative learning model based on the Mask R-CNN instance segmentation method (He et al. 2017). Collaborative learning entails the inclusion of two or more deep learning models to achieve superior results than can be obtained using a single model. In our system, two models were independently trained and created: one for tooth segmentation and another for filling segmentation. The outputs of the two models are combined to form a single image to identify and classify teeth. By combining the outcomes of different models, the collaborative model can provide a summary from the inferences of multiple tasks, resulting in superior accuracy. Our present model consists of two steps: (1) deep learning modeling and inferencing using two individual models (primary and permanent tooth segmentation and filling segmentation) and (2) inference aggregation for multiple tasks, providing a summary of the inferencing outcomes to generate a comprehensive understanding of the tooth locations, types, and the presence of fillings.

## Methods

### Datasets

The primary dataset used for tooth and filling segmentation was the Universidade Federal Da Bahia-Universidade Estadual de Santa Cruz (UFBA-UESC) dental dataset (Silva et al. 2018), which included 368 panoramic radiographs used to train the tooth and filling segmentation models. We also used 80 deidentified panoramic radiographs from the University of Missouri-Kansas City (UMKC) School of Dentistry. The details of the total number of panoramic radiographs used for training, validation and testing from the datasets are shown in Table 1, as well as the number of panoramic radiographs with fillings. The analysis of these deidentified panoramic radiographs was determined to be exempt by the Institutional Review Board of UMKC (IRB Project Number 2068642; IRB Review Number 334839).

**Table 1.** The datasets used in the present study

|  | UFBA-UESC |  |  | UMKC |  |  |
| --- | --- | --- | --- | --- | --- | --- |
|  | *Training* | *Validation* | *Testing* | *Training* | *Validation* | *Testing* |
| Number of Panoramic Radiographs | 368 | 246 | 65 | 80 | 29 | 35 |
| Number with Fillings | 117 | 60 | 65 | 70 | 40 | 35 |

UFBA-UESC: Universidade Federal Da Bahia-Universidade Estadual de Santa Cruz (UFBA-UESC) dental dataset described by (Silva et al. 2018)
UMKC: University of Missouri-Kansas City

### Modeling and Inferencing

We first developed two distinct models for the segmentation of primary and permanent teeth and for filling segmentation using panoramic radiographs (Figure 1A). Then we performed inference utilizing these models, and the results were forwarded to the consequence phase for aggregation.

**Figure 1:**
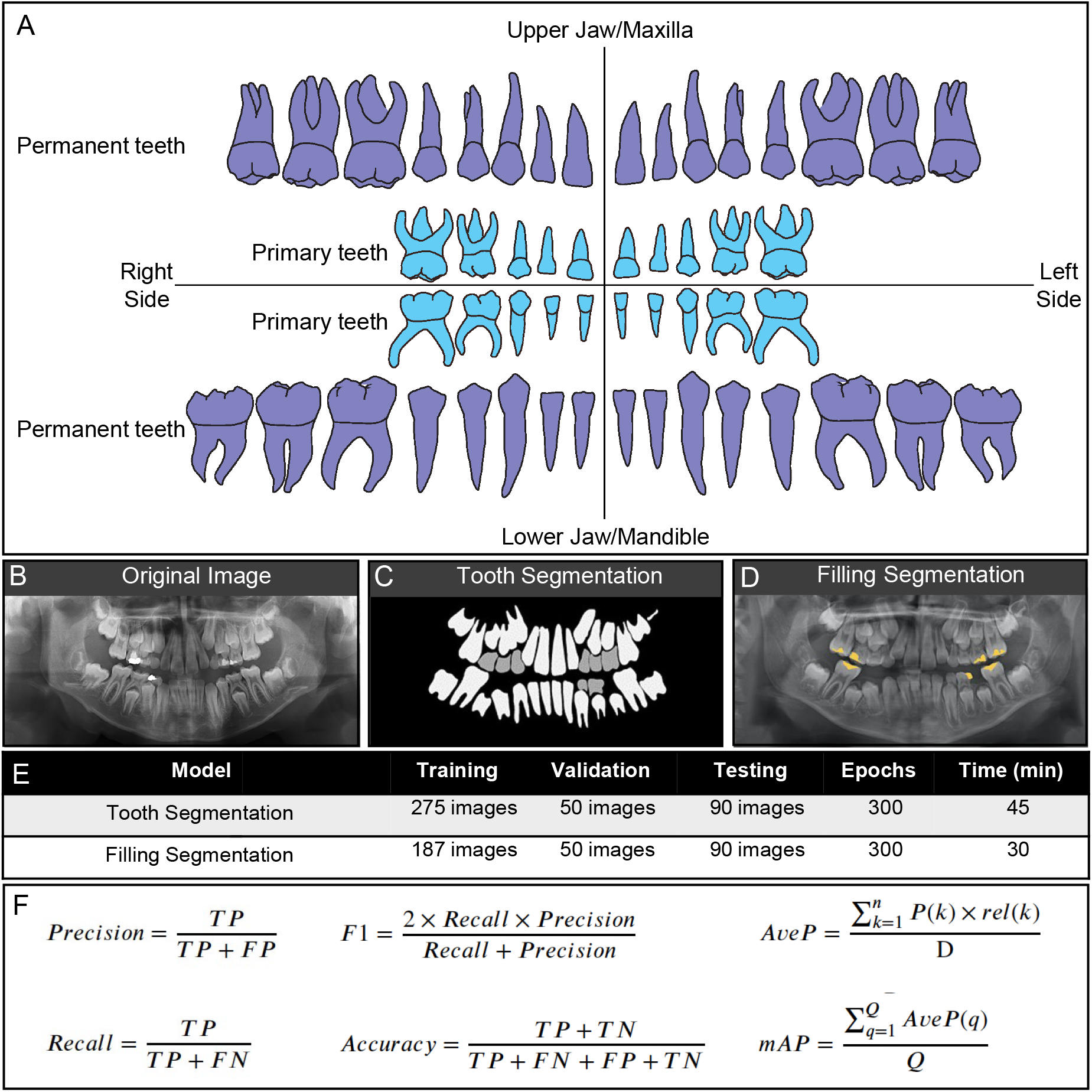
An Overview of the Models. **(A)** The panoramic appearance of the full primary dentition (light blue) and permanent dentition (purple) is shown. **(B)** A panoramic radiograph of a patient with mixed dentition, with fillings present in both the primary and permanent teeth. **(C)** The tooth segmentation mask of the panoramic radiograph in (B) with permanent teeth segmented in white and primary teeth segmented in grey. **(D)** The filling segmentation mask overlaying the panoramic radiograph shown in (B). **(E)** An overview of the tooth segmentation and filling segmentation models, including the panoramic radiographs used, number of images used for training, testing and validation, as well as the Epoch number and time. **(F)** The equations used to determine the precision, recall, F1 score, accuracy, average precision measure (AveP) and mean average precision (mAP). TP denotes true positive (correct segmentation), TN denotes true negative, FP denotes false positive, and FN denotes false negative. Precision was assessed as the proportion of positive class predictions (TP+FP) that were genuinely positive (TP). Recall quantifies the number of positive class predictions (TP) made from the dataset’s positive examples (TP+FN). The F1 score accounts for both precision and recall concerns. The average precision measure (AveP) was calculated as noted in Figure 1F, where D denotes the total number of relevant documents and rel(k) denotes an indicator function equal to one if the item at rank k is a relevant document (and is set as zero otherwise). The mAP is calculated as the average of the precision scores for each query, where Q is the total number of inquiries.

### Model 1: Primary and Permanent Tooth Segmentation Modeling

To segment teeth, we utilized the Mask R-CNN with the UFBA-UESC & UMKC panoramic radiograph datasets. This instance-based segmentation model can handle both segmentation and two-category identifications, and is able to differentiate between primary and permanent teeth. Figure 1B shows an example of a panoramic radiograph from a patient with mixed dentition and fillings. The tooth segmentation mask for the panoramic radiograph analyzed by our model is annotated for the segmentation of 8 primary and 32 permanent teeth (Fig. 1C). In the model, Mask R-CNN retrieves features from ResNet-101, which performs feature extraction. Subsequently, a feature pyramid network (FPN) with anchors is formed utilizing the regions of interest (ROIs) identified. After aligning the ROIs, the classification and localization of all teeth are carried out based on the regression of the bounding boxes for teeth. Finally, the convolutional network is utilized to identify and segment each tooth, as indicated by the bounding boxes. The tooth segmentation model was created using the panoramic radiograph datasets and Facebook Research’s Detectron2 Library for python 3.7 (Li et al. 2019, Wu et al. 2019).

### Model 2: Filling Segmentation Modeling

Deep CNN methodologies are among the most effective and practical methods for identifying fillings. We used the Mask R-CNN (initially described by He et al. 2017) for filling segmentation. Dental panoramic radiographs were manually annotated to identify fillings and were used for segmentation learning for detection. The filling segmentation mask for the panoramic radiograph shown in Figure 1B was analyzed by our model, and was annotated for 7 fillings (Fig. 1D).

Individual detection models for tooth segmentation and filling segmentation were developed separately by using the benchmark dataset from the UFBA-UESC (Silva et al. 2018). Training the tooth segmentation model with the panoramic radiographs required around 448 annotated training images, 275 for validation and 100 testing images with primary and permanent teeth (Fig. 1E). For filling segmentation, 187 images were used for training, 100 for validation, and 100 were used for testing the model (Fig. 1E). All types of fillings were annotated (amalgam, composite, glass ionomer etc.), but other types of restorations were not annotated for this analysis (implants, crowns, etc.). Around 100 pediatric images were utilized to evaluate the collaborative model, which was developed by combining the output from the two distinct segmentation models.

### Collaborative Learning

The collaborative model’s initial stage involves concurrent inference from the two models, followed by aggregation of the predictions of these models, and finally the generation of a summary of the results. The present collaborative model’s novel design allows it to draw inferences from multiple models using a single input for a variety of relevant tasks, in this case, tooth segmentation and filling segmentation. It then provides a summary of data from the different models. A collaboration model is capable of doing multiple tasks with a low dependence on the individual models selected. At this stage, the accuracy is determined as the weighted average of the accuracy of the distinct models. The initial weights are equal for each model, but the weight can be changed after analyzing the contributions of each individual model. The integrated inference summary is saved in a standard format (Microsoft COCO) that can be used for further analyses.

### Evaluation Measures

In the present study, we evaluated both individual models (tooth segmentation with primary versus permanent identification and filling segmentation) as well as collaborative models. We employed a variety of metrics to assess the performance of the models, including the accuracy, precision, recall, performance (F1 score), and mean average precision (mAP) (Fig. 1F).

## Results

### Collaborative Model Derived from the Tooth and Filling Segmentation Models

Collaborative inference was performed by combining the outputs of the two segmentation models (see Fig. 1). We validated our findings using images that were not included in the training data. After detecting and labeling teeth, an performance accuracy of approximately 95% was obtained for the tooth segmentation, including the identification of primary versus permanent teeth (Table 2). Individual cases illustrating the efficacy of the model are described below.

**Table 2.** The performance of the individual and collaborative models for tooth and filling segmentation

|  | Average number |  |  | Tooth Segmentation Model |  | Filling Segmentation Model |  | Collaborative Model |  |
| --- | --- | --- | --- | --- | --- | --- | --- | --- | --- |
|  | *Primary teeth* | *Permanent teeth* | *Fillings* | *mAP* | *F-1* | *mAP* | *F-1* | *mAP* | *F-1* |
| Overall | 1264 (# of teeth) | 8593 (# of teeth) | 2490(# of filling) | 95.32% | 92.50% | 91.53% | 91% | 94.09% | 93.41% |
| Example Case 1 (shown in Fig. 2) | 32 | 0 | 0 | 98.15% | 98.63% | 99.45% | 99.74% | 98.86% | 99.18% |
| Example Case 2 (shown in Fig. 3) | 32 | 0 | 8 | 98.67% | 98.75% | 99.33% | 99.65% | 99.04% | 99.28% |
| Example Case 3 (shown in Fig. 4) | 32 | 8 | 0 | 97.89% | 98.45% | 99.57% | 99.67% | 99.03% | 99.30% |
| Example Case 4 (shown in Fig. 5) | 32 | 9 | 5 | 97.54% | 98.15% | 99.24% | 99.54% | 98.67% | 99.07% |

### Case 1: Permanent Dentition

A panoramic radiograph of a patient with only permanent dentition is shown in Fig. 2A. The tooth segmentation model successfully identified all 32 permanent teeth in the tooth segmentation mask (Fig. 2B). The filling segmentation model confirmed that there were no fillings in the filling segmentation mask (Fig. 2C). The collaborative model had an accuracy of 98.86% for this case (Fig. 2D & E). The F-1 score and mAP for this patient are shown in Table 2.

**Figure 2:**
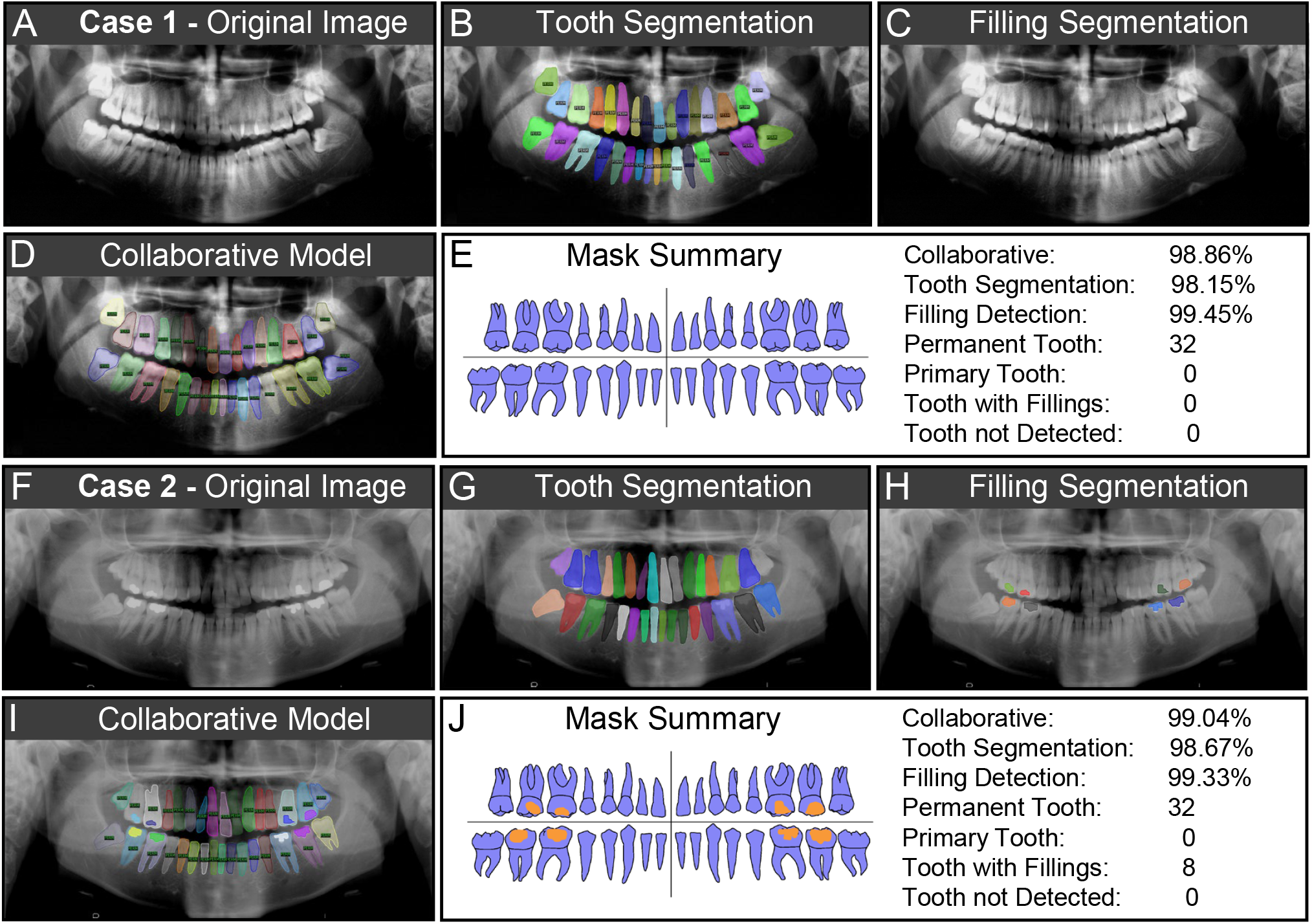
Example Cases of Permanent Dentition and Permanent Dentition with Fillings. Case 1: **(A)** The original panoramic radiograph of a patient with only permanent dentition. **(B)** The tooth segmentation mask overlaying the original panoramic radiograph. **(C)** The filling segmentation mask overlaying the original panoramic radiograph – no fillings were noted. **(D)** The collaborative model mask overlaying the original panoramic radiograph. **(E)** A mask summary showing that all 32 permanent teeth were identified (shown in purple) and listing the accuracies of the models. **Case 2: (F)** The original panoramic radiograph of a patient with only permanent dentition and multiple fillings. **(G)** The tooth segmentation mask overlaying the original panoramic radiograph. **(H)** The filling segmentation mask overlaying the original panoramic radiograph – four fillings were noted. **(I)** The collaborative model mask overlaying the original panoramic radiograph. **(J)** A mask summary showing all 32 permanent teeth (purple) with 4 fillings (identified in orange), and listing the accuracies of the models.

### Case 2: Permanent Dentition with Fillings

Figure 2F shows a panoramic radiograph from a patient with only permanent dentition present and fillings in the permanent dentition. The tooth segmentation model successfully identified all 32 permanent teeth in the tooth segmentation mask (Fig. 2G). The filling segmentation model identified all 8 fillings in the filling segmentation mask (Fig. 2H). The collaborative model showed an accuracy of 99.04% for this case (Fig. 2I & J). The corresponding F-1 score and mAP are also shown in Table 2.

### Case 3: Mixed Dentition

Case 3 examined a panoramic radiograph from a patient with a mixed dentition, where both primary and permanent teeth were present (Fig. 3A). The tooth segmentation model could identify all 32 permanent teeth and 8 primary teeth in the tooth segmentation mask (Fig. 3B). The filling segmentation model confirmed that there were no fillings in the filling segmentation mask (Fig. 3C). The collaborative model showed an accuracy of 99.03% for this case (Fig. 3D & E). The corresponding F-1 score and mAP are shown in Table 2.

**Figure 3:**
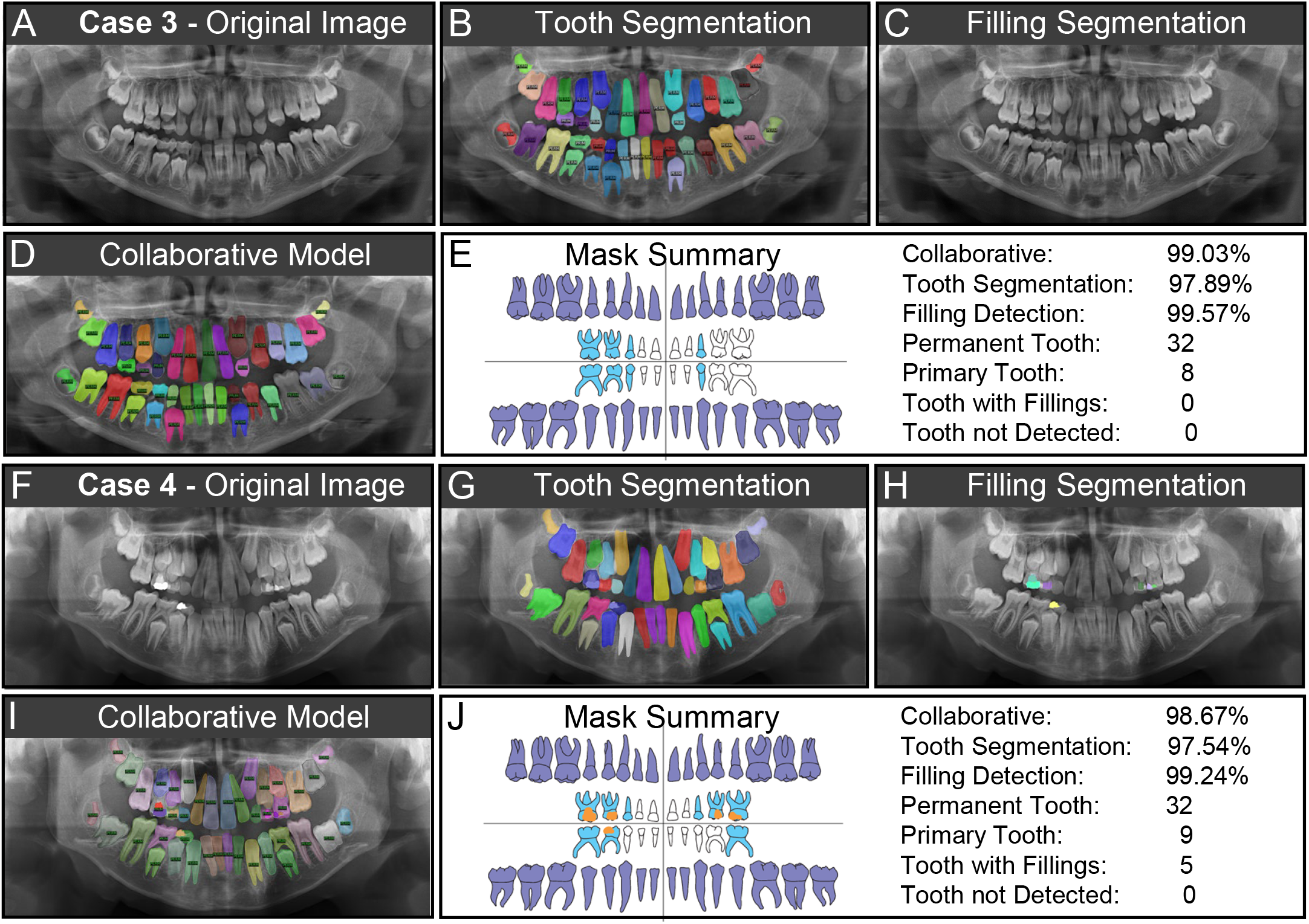
Example Cases of Mixed Dentition and Mixed Dentition with Fillings. Case 3: **(A)** The original panoramic radiograph of a patient with mixed (primary + permanent) dentition. **(B)** The tooth segmentation mask overlaying the original panoramic radiograph. **(C)** The filling segmentation mask overlaying the original panoramic radiograph – no fillings were noted. **(D)** The collaborative model mask overlaying the original panoramic radiograph. **(E)** A mask summary showing all 32 permanent teeth (purple) and 8 primary teeth (blue). The accuracies of the models are also shown. **Case 4: (F)** The original panoramic radiograph of a patient with a mixed dentition with multiple fillings. **(G)** The tooth segmentation mask overlaying the original panoramic radiograph. **(H)** The filling segmentation mask overlaying the original panoramic radiograph – five fillings were noted. **(I)** The collaborative model mask overlaying the original panoramic radiograph. **(J)** A mask summary showing all 32 permanent teeth (purple), 9 primary teeth (blue), and 5 fillings (orange), in addition to the accuracies of the models.

### Case 4: Mixed Dentition with Fillings

A panoramic radiograph from a patient with mixed dentition and fillings in the primary dentition is shown in Fig. 4F. The tooth segmentation identified all 32 permanent teeth and 9 primary teeth in the tooth segmentation mask (Fig. 4G). The filling segmentation model identified all 5 fillings in the primary teeth in the filling segmentation mask (Fig. 4H). Finally, the collaborative model showed an accuracy of 98.67% for this case (Fig. 4I & J). The corresponding F-1 score and mAP are shown Table 2.

## Discussion

The accuracy of our present model was comparable to (or superior to) that of the previous tooth segmentation approaches for panoramic radiographs. Moreover, we enhanced the F1 score from an average of 92.5% for the tooth segmentation model alone to 93.41% for the collaborative model (Table 2). We are currently working to enhance the performance of our collaborative learning model by optimizing the models’ inferencing and weighting functions, as well as improving collaboration strategies.

Our proposed model is distinct from existing models in that it executes multiple tasks and generates a summary of each individual model’s findings. There have only been a few studies that have segmented primary teeth, and most of these have focused on identifying specific abnormalities (e.g., mesiodens) (Ahn et al. 2021, Ha et al. 2021, Jeon et al. 2022, Kaya et al. 2022, Kılıc et al. 2021, Pinheiro et al. 2021). Moreover, while various deep learning methods have been developed to detect fillings, to our knowledge, the present model is the first to detect fillings in both primary and permanent teeth. Our present method also performed as well as or better than the previous deep learning models for tooth and filling segmentation by providing the collaborative model (Cui et al. 2019 and 2021, Estai et al. 2022, Lee et al. 2020, Leite et al. 2021, Shaheen et al. 2021, Silva et al. 2020, Vinayahalingam et al. 2021, Wirtz et al. 2018, Zhang et al. 2020, Zhao et al. 2020).

Nevertheless, there are several limitations associated with our present method. First, the data presented here were from a relatively small number of patients, and the patient images were from only two geographic regions (Brazil and central USA). Additional training of the model using more patient images will be undertaken in future studies. As noted above, we are currently working to further improve the accuracy of the model by addressing the relative importance of the different segmentation data and including lower-quality images, as well as images from patients with abnormal dentition. We are also working to include the identification of specific tooth numbers to improve the segmentation and classification of the teeth (Chandrashekar et al. 2022). Finally, while the present study included various types of fillings, no other dental restorations were evaluated. A more comprehensive model including a wider range of variables would be useful for clinical practice.

Although more work is needed, the present collaborative model is able to simultaneously segment primary and permanent teeth, as well as any fillings present, with high accuracy. The support of systems using artificial intelligence and machine learning can help dentists ensure that they are providing the best care to their patients.

## Data Availability

All data produced in the present study are available upon reasonable request to the authors.

## Author Contributions

E. Bumann contributed to the conception, design, data acquisition and interpretation, and also drafted and critically revised the manuscript. S. Al-Qarni contributed to the conception, design, data acquisition and interpretation, performed statistical analyses, and critically revised the manuscript. G. Chandrashekar contributed to the conception, design, data acquisition and interpretation, performed statistical analyses, and critically revised the manuscript. R. Sabzian contributed to the design, data acquisition and interpretation, and critically revised the manuscript. B. Bohaty contributed to the conception and design, and critically revised the manuscript. Y. Lee contributed to the conception, design, data acquisition and interpretation, and critically revised the manuscript. All authors gave their final approval and agreed to be accountable for all aspects of the work.

